# Exploratory functional and quality of life outcomes with daily consumption of the ketone ester bis-octanoyl (R)-1,3-butanediol in healthy older adults: a randomized, parallel arm, double-blind, placebo-controlled study

**DOI:** 10.1101/2024.09.17.24313811

**Authors:** Brianna J Stubbs, Elizabeth B Stephens, Chatura Senadheera, Stephanie Roa Diaz, Sawyer Peralta, Laura Alexander, Wendie Silverman-Martin, Jamie Kurtzig, B. Ashen Fernando, James T Yurkovich, Thelma Y Garcia, Michi Yukawa, Jennifer Morris, James B Johnson, John C Newman

## Abstract

**Background:** Ketone bodies are metabolites produced during fasting or on a ketogenic diet that have pleiotropic effects on the inflammatory and metabolic aging pathways underpinning frailty in *in vivo* models. Ketone esters (KEs) are compounds that induce hyperketonemia without dietary changes and that may impact physical and cognitive function in young adults. The functional effects of KEs have not been studied in older adults.

**Objectives:** Our long-term goal is to examine if KEs modulate aging biology mechanisms and clinical outcomes relevant to frailty in older adults. Here, we report the exploratory functional and quality-of-life outcome measures collected during a 12-week safety and tolerability study of KE (NCT05585762).

**Design:** Randomized, placebo-controlled, double-blinded, parallel-group, pilot trial of 12-weeks of daily KE ingestion.

**Setting:** The Clinical Research Unit at the Buck Institute for Research on Aging, California. Participants: Community-dwelling older adults (≥ 65 years), independent in activities of daily living, with no unstable acute medical conditions (n = 30).

**Intervention:** Subjects were randomly allocated (1:1) to consume 25 g daily of either KE (bis-octanoyl (R)-1,3-butanediol) or a taste, appearance, and calorie-matched placebo (PLA) containing canola oil.

**Measurements:** Longitudinal change in physical function, cognitive function and quality of life were assessed as exploratory outcomes in n = 23 completers (n = 11 PLA, n = 12 KE). A composite functional outcome to describe the vigor-frailty continuum was calculated. Heart rate and activity was measured throughout the study using digital wearables.

**Results:** There were no statistically significant longitudinal differences between groups in exploratory functional, activity-based or quality of life outcomes.

**Conclusion:** Daily ingestion of 25 g of KE did not affect exploratory functional or quality-of-life end points in this pilot cohort of healthy older adults. Future work will address these endpoints as primary and secondary outcomes in a larger trial of pre-frail older adults.

## Introduction

The Geroscience Hypothesis, though yet untested in clinical trials, postulates that intervening in common aging biological mechanisms could mitigate or prevent various chronic age-related diseases as well as geriatric syndromes such as frailty (1). Frailty syndrome, characterized by diminished physiological reserves and heightened susceptibility to adverse health stressors, posing a significant risk for disability, institutionalization, and mortality (2). The prevalence of frailty escalates with age, with around 15% of adults over 65 years old in the United States meeting one standard definition (3). Although the pathophysiology of frailty is multi-system and poorly elucidated, it is believed to encompass deficits in cellular energy production, chronic inflammation, and immune dysfunction (4, 5) – all of which also comprise geroscience molecular mechanisms commonly recognized as the "Hallmarks of Aging" (6). Presently, specific molecular therapies for frailty remain elusive, although interventions such as exercise programs, specialized geriatric care models, and nutritional strategies have shown the most promise in modulating the frailty phenotype and its sequalae (7).

A challenge in interventional studies of frailty is the choice of a primary outcome to capture clinically meaningful changes in function, especially as many outcomes focus on the frailest population, and poorly discriminate among those ‘not frail’. Recently, Newman *et al.*, (8) used a large (n = ∼900), well-characterized older adult population to develop a four-item continuous composite score (comprising digit symbol substitution, leg power, peak oxygen consumption and fatigability) that better captures the spectrum of function from vigorous to frail. The score was strongly associated with age, with lower scores predicting functional limitation (8). Such composite scores both increase the chance of detecting intervention-driven differences in higher functioning people and avoid the selection of a single arbitrary outcome for a complex, multi-factorial condition such as frailty and pleiotropic interventions such as diet or exercise.

An increasing number of studies have explored the impact of dietary restriction and fasting on aging biology in animal models (9) and humans (e.g., CALERIE (10), HALLO-P (NCT05424042)). One key shared feature of dietary restriction and fasting is the presence of ketone bodies, notably beta-hydroxybutyrate (BHB), synthesized in the liver during fasting or carbohydrate restriction. The primary role of BHB is to provide an alternative energy source to various tissues, including the brain, muscle, and heart. The energetic and molecular signaling activities of ketone bodies, including improving mitochondrial function and regulating inflammatory activation, support a mechanistic role in modulating aging and may be directly relevant to frailty (11). Prior studies have demonstrated the potential of increasing circulating ketone concentrations in extending healthy lifespan and ameliorating age-related functional decline in animal models (12, 13).

Ketone esters (KEs), such as bis-octanoyl (R)-1,3-butanediol, are examples of exogenous ketones, small molecules that deliver ketone bodies without other dietary changes (14, 15). KEs are hydrolyzed in the gut to release ketogenic precursors, which are then metabolized in the liver to release ketone bodies (16). In preclinical models, ketone bodies and KEs attenuate muscle atrophy through anticatabolic signaling activities (17), improve heart function in age-related heart failure (18, 19), and promote healthy function of T cell subpopulations (20, 21). Clinically demonstrated effects of acute KE administration range from blood glucose control (22, 23) to physical (24, 25), cognitive (26, 27), immune (28), and cardiovascular (27, 29) function in younger adult populations under the age of 65. These observations have led to our central hypothesis, that ketone bodies delivered through KEs may ameliorate the frailty syndrome through multi-system energetic and signaling activities that improve metabolic and immune function. To date, there are no published studies describing the effects of KE on functional outcomes in an aging population.

To begin addressing this gap, and laying the foundation for future work testing the KE in a pre-frail and frail population (NIH funding ID: **1R01AG081226-01)**, we undertook a 12-week randomized, placebo-controlled, double-blinded, parallel-group pilot clinical trial with the primary objective of generating the first long-term safety and tolerance data for KEs in an independent living, generally healthy population of older adults. Here we report data describing our exploratory aging-focused functional endpoints. We also calculated a four-item composite outcome score based on the work of *Newman et al (8)* to investigate functional changes on a continuum from vigorous to frail. Based on the positive data of short-term KE use in younger adults, we hypothesized that 12-weeks of KE consumption would improve functional outcomes in our older adult population.

## Methods

### Study Design Overview

Healthy older adults ≥ 65 years of age (n = 30) took part in this randomized, 12-week randomized, placebo-controlled, double-blinded, parallel-group, pilot clinical trial (Figure 1). The full study design was powered to determine tolerance and safety and has been previously described (30, 31). The study included single-administration kinetics visits prior to starting and after completing the main 12-week protocol, results of which are reported separately (32). Physical, cognitive, and quality of life (QoL) exploratory outcomes were measured at week 0 and week 12. Participants arrived at these visits fasted for blood draws, then were provided with a snack and, if desired, coffee or tea prior to beginning functional testing. Wrist based health and fitness monitors were distributed at the first kinetics visit (at least 7 days before baseline) and were worn continuously until the final visit. The study was approved by Advarra IRB on September 28^th^ 2022 (Pro00065464). The first subject was randomized on January 31^st^ 2023 and the final subject completed the trial on January 17^th^ 2024, which ended the study. The study was conducted in accordance with the 2013 Declaration of Helsinki.

**Figure 1:**
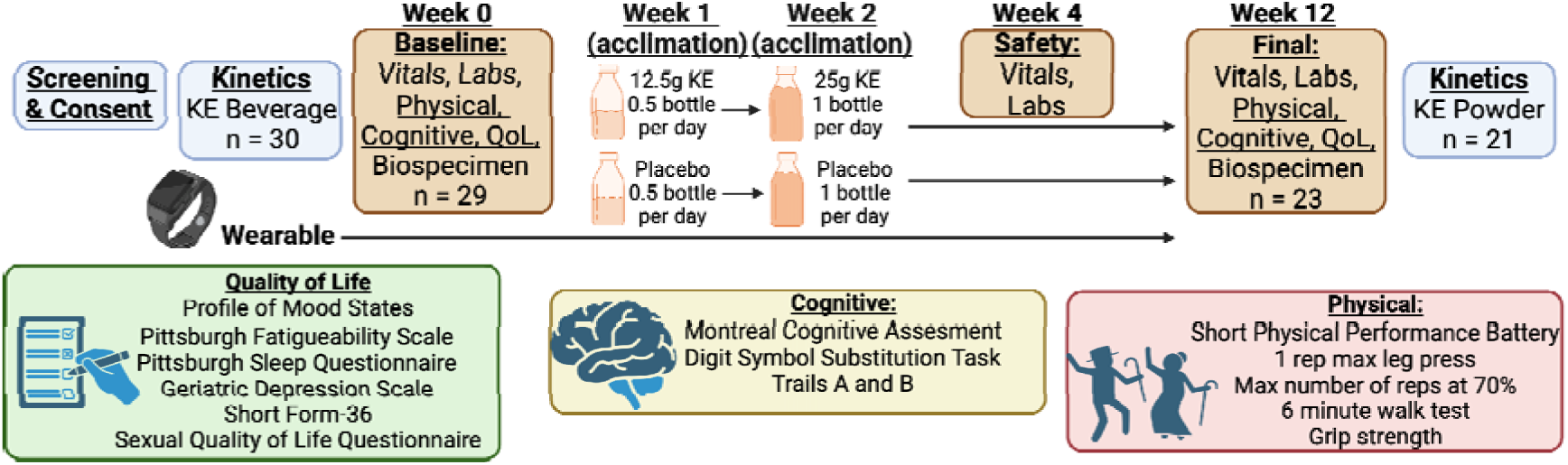
Study schematic showing the schedule of visits and assessments described in this article. **Abbreviations:** KE, ketone ester; QoL, quality of life. Created using BioRender.com

### Participants

Participants were community dwelling older adults (≥ 65 years of age), independent in activities of daily living and in stable health. The full inclusion and exclusion criteria are listed in the **Supplemental Information**. Decisions on eligibility were made by an independent medical officer to ensure allocation was concealed, and eligible participants were randomized by the study team based on a statistician-generated block allocation sequence (block size 4, intended to equally randomize male and female participants). The flow of study participants is illustrated in the CONSORT diagram (**Supplementary Figure 1**). Sample size was determined *a priori* based on the primary outcome of the study, which was tolerance of the KE beverage (30), and feasibility of this pilot.

### Study Beverages

The study KE beverage was a tropical-flavored beverage containing the KE bis-octanoyl (R)- 1,3-butanediol (Cognitive Switch, BHB Therapeutics Ltd, Dublin, IRE). Each bottle was 75 mL and contained 25 g of KE. Participants were instructed to consume the study beverage at home daily for 12 weeks within 5 minutes of their first meal of the day. Half of a bottle (12.5 g KE) was consumed daily during week 1, and a full bottle (25 g KE) for the remainder of the study. The placebo used in the study was custom manufactured by BHB Therapeutics. In the placebo the KE was replaced with a non-ketogenic canola oil and matched for volume, appearance, flavor and calories. Nutritional facts for study beverages are shown in **Supplemental Table 2**. Beverages were provided as single serving bottles and labeled with the coded group allocation. All personnel involved with the data collection, subject assessment, analysis, and interpretation were blinded to the identity of the intervention assigned to participants, but not the allocation.

### Frailty Indices

All frailty indices were evaluated by self-report during an interview in the screening and final visits as follows: Katz Index of Independence in Activities of Daily Living (Katz ADL) (33), Lawton Instrumental Activities of Daily Living (Lawton IADL) (34), and Canadian Study of Health and Aging (CSHA) Clinical Frailty Scale (CFS) (35, 36).

### Physical Function

Physical function measures were collected at the baseline and final visits as follows: 1 repetition maximum (1-RM) leg press strength, 70% of 1-RM leg press fatiguability (both using Matrix Versa, Matrix Fitness, Cottage Grove, USA), Short Physical Performance Battery (SPPB), 4 m gait speed (as part of the SPPB), 6-minute walk test, grip strength, and actigraphy (FitBit Inspire 2, FitBit, San Francisco, USA). Subjects were familiarized with the 1-RM equipment during the first kinetics visit.

#### One Repetition Maximum (1-RM) Leg Press Strength

Subjects were instructed on proper positioning and form on the seated leg press machine. They were positioned with their back straight and against the back rest, their feet on the footplate so that knee and hip angles were approximately 90°and the weight sled was adjusted accordingly. Foot position and sled position were replicated for both visits. The Rating of Perceived Exertion (RPE) Category-Ratio Scale was used to measure effort (37), RPE of 9-10 was the target. First, subjects performed 5 - 6 unweighted repetitions as familiarization and to correct positioning or form. Then, subjects completed a decreasing number of repetitions (starting repetitions = 6 - 7) at a set weight (starting weight = 50% body weight) and rated their exertion after each set. Weight increased and number of repetitions decreased incrementally until the target RPE was achieved and the participant could only perform one repetition. Standardized rest was timed between each set and participants were verbally encouraged throughout the test.

#### Leg Press Fatiguability

Leg press fatiguability was performed after the 1-RM leg press after at least 5 minutes of rest. Positioning was replicated from the 1-RM leg press strength test. Weight was set to 70% of the 1-RM and subjects were instructed to complete as many repetitions as possible whilst maintaining proper form and a consistent cadence. Improper form was corrected once, the test was ended if a second correction was required. Exertion was rated using the RPE Category-Ratio Scale (37) immediately following completion. Participants were given verbal encouragement throughout the test.

#### Short Physical Performance Battery (SPPB)

The SPPB includes several short tests to evaluate lower extremity function, including measures of standing balance, 4-meter gait speed, and ability to rise from a chair 5 times, and was administered according to standard instructions (38).

#### 6-Minute Walk Test

A 10-meter-long course was measured and marked with tape at each meter interval and a cone at each end. Subjects were instructed to walk briskly from cone to cone, rounding the cones to invert direction, for 6 minutes. Subjects were provided with standard encouragement throughout. Before walking began and immediately following completion, subjects measured their exertion and breathing difficulty using the RPE Category-Ratio Scale and Modified Borg Dyspnea Scale (37). After 6 minutes, subjects stopped in place, and partial laps were recorded.

#### Grip Strength

Grip strength was measured using a Jamar Hydraulic Hand Dynamometer (Jamar, Warrenville, IL, USA). Participants were sat in an armed chair and instructed to rest their arms on the arm rests with their elbow at a 90° angle. The dynamometer handgrip was adjusted to a setting comfortable for the subject, and a submaximal practice test was performed for familiarization. Three trials were performed per hand, and hands were alternated between trials. Measurements were recorded in kgs and rounded to the nearest 2 kg.

### Health and Fitness Tracker

Wrist health and fitness trackers were dispensed at the kinetics visit at least 7 days before the baseline visit. Subjects were instructed to wear the device throughout the 12-week trial, except for charging. The device was used to record heart rate, steps, sleep (total time asleep and sleep efficiency), and activity (sedentary, lightly active, moderately active, and very active minutes).

### Cognitive Function

Cognitive function measures were collected at the baseline and final visits as follows: Montreal Cognitive Assessment (MoCA), Trails Test A and B, and Digit Symbol Substitution Test (DSST). All cognitive function tests were conducted by trained study staff according to standard protocols, in a private, quiet room. The MoCA was scored by a study physician.

### Quality of Life

QoL measures were collected via self-rated questionnaire at the baseline and final visits as follows: Geriatric Depression Scale (GDS), Pittsburgh Sleep Quality Index (PSQI), 36-Item Short Form Health Survey (SF-36, Version 1, Rand), Profile of Mood States—Short Form (POMS-SF), sexual quality of life questionnaire (SQoL), Pittsburgh Fatigability Scale (PFS). All quality-of-life questionnaires were conducted on paper with pen in a private room. Questionnaires were scored by trained study staff.

### General Statistical Design

Statistical anaysis was carried out using Prism 10, except for actigraphy analysis, which was carried out in R and Python. Data is shown as mean (standard deviation), unless otherwise specified. Significance was set at p < 0.05. P values are not adjusted for multiple comparisons given the exploratory nature of these analyses.

To analyze individual physical, cognitive and QoL outcomes, we firstly confirmed there were no differences at baseline between intervention groups; raw baseline values for KE and PLA group were compared using an unpaired t-test or a Mann-Whitney Test, as appropriate. If there was no significant difference between groups at baseline, a Week 0 to 12 change value was calculated for each outcome in each individual who completed the study per protocol. The between-group difference in change value was then compared using an unpaired t-test or a Mann-Whitney Test, as appropriate.

### Composite Frailty Score

A composite frailty score was calculated based on the rationale of Newman *et al* (8). We included four outcomes: leg strength (1 rep max leg press weight), endurance (6 minute walk test), speed (digit symbol substitution) and perceived Fatigability (Pittsburgh Fatigability Scale – Physical). A z-score for each individual at baseline was calculated, and a z-score for the pre- to post- change in per protocol subjects was calculated using the standard deviation of the pooled baseline values. Z-scores were calculated as sex-specific. The z-scores of the four items of interest were averaged to give the final composite score. A simple linear regression was calculated for the the baseline composite score vs. age and the pre- to post- change in composite score vs. age.

### Health and Fitness Tracker Data Analysis

Health and fitness tracker data was analyzed seperately for each variable of interest, data was compiled for all per protocol subjects and duplicates were removed. Heart rate data was processed by filtering out low confidence measurements (confidence < 2). We determined sleep timing through the Fitbit’s sleep staging, and mapped these time windows with the heart rate data to determine sleeping heart rate. We then calculated the sleeping heart rate (SHR) to be the mean value over each night-time sleep period for each individual. Time asleep was determined by the Ftibit’s sleep staging, excluding all daytime naps (one dataset was missing). Minutes of sedentary, light, moderate and vigorous activity was determined through Fitbit’s Active Minutes function that combines heart rate and movement data. Total minutes of each activity type per day was calculated by adding all data points per date. All data was analyzed by plotting the mean of each value per allocation, with bands generated using the standard error. Additional figures plotted each individual’s time course with an aggregate mean value per allocation. These data were analyzed using R 4.3.1 and Python.

## Results

### Participants and completion

A total of 30 participants were randomized (mean age (range), Male n = 15; age = 76.5 y (65 – 90); mean BMI (range) = 25.2 (19.9 – 32.7); median Katz ADL (range) = 6 (5-6), median Lawton IADL (range) = 8 (8-8), median CSHA Frailty Score (range)= 1 (1 – 3)). Full participant anthropometric characteristics and frailty indices at baseline are shown in **Supplemental Table 1**.

Subject disposition is shown in a CONSORT diagram (**Supplemental Figure 1**). Briefly, 23 subjects completed the 12-week protocol (the per-protocol population), 1 participant completed the acute kinetics visit but did not start the main study, without giving a reason, and 6 participants dropped out after Day 0 and did not complete the full protocol; repeat functional data was not available for subjects who did not start or finish the study. Adherence with consumption of the study products for the per protocol population was high, with n = 15 reporting 100% adherence and a range of 94 – 99 % for the remaining 8 participants who completed the study, based on study logs and returned bottles.

### Frailty Indicies and Frailty-Vigor Composite

No subjects experienced a change in Katz ADL or Lawton IADL scores after the intervention. The CSHA Frailty Score was changed by one point in n = 3 subjects (PLA = 2, one increase, one decrease, KE = 1, increase). There were no significant differences between groups in pre- to post- change in any of the individual items included in the composite score (**Figure 2 A-D)**: 1 rep max leg pres (KE = -0.8 (13.4) kg, PLA = 0.9 (10.9) kg, p = 0.834), Six Minute Walk Test (KE = 42.1 (48.1) m; PLA = 30.9 (38.8) m, p = 0.705), Digit Symbol Substitution Task (correct) (KE = 2.4 (4.6); PLA = 2.6 (4.0), p = 0.706), Pittsburgh Fatigability Scale (Physical) (KE = 0.1 (3.6); PLA = -1.6 (5.2), p = 0.711). When the items were converted to z-scores and combined into the 4-item composite outcome, there was no significant difference between groups (KE = 0.21 (0.25); PLA = 0.17 (0.23), p = 0.833, **Figure 2E**). The baseline composite score trended down with increasing age (F = 2.037; df = 1, 27; P = 0.165; R^2^ = 0.07; **Figure 2F**), and the change in composite score over the 12 week intervention was not related to the age of the subject (F = 0.002; df = 1, 21; P = 0.967; R^2^ = 8.26 x 10^-5^); **Figure 2G**).

**Figure 2:**
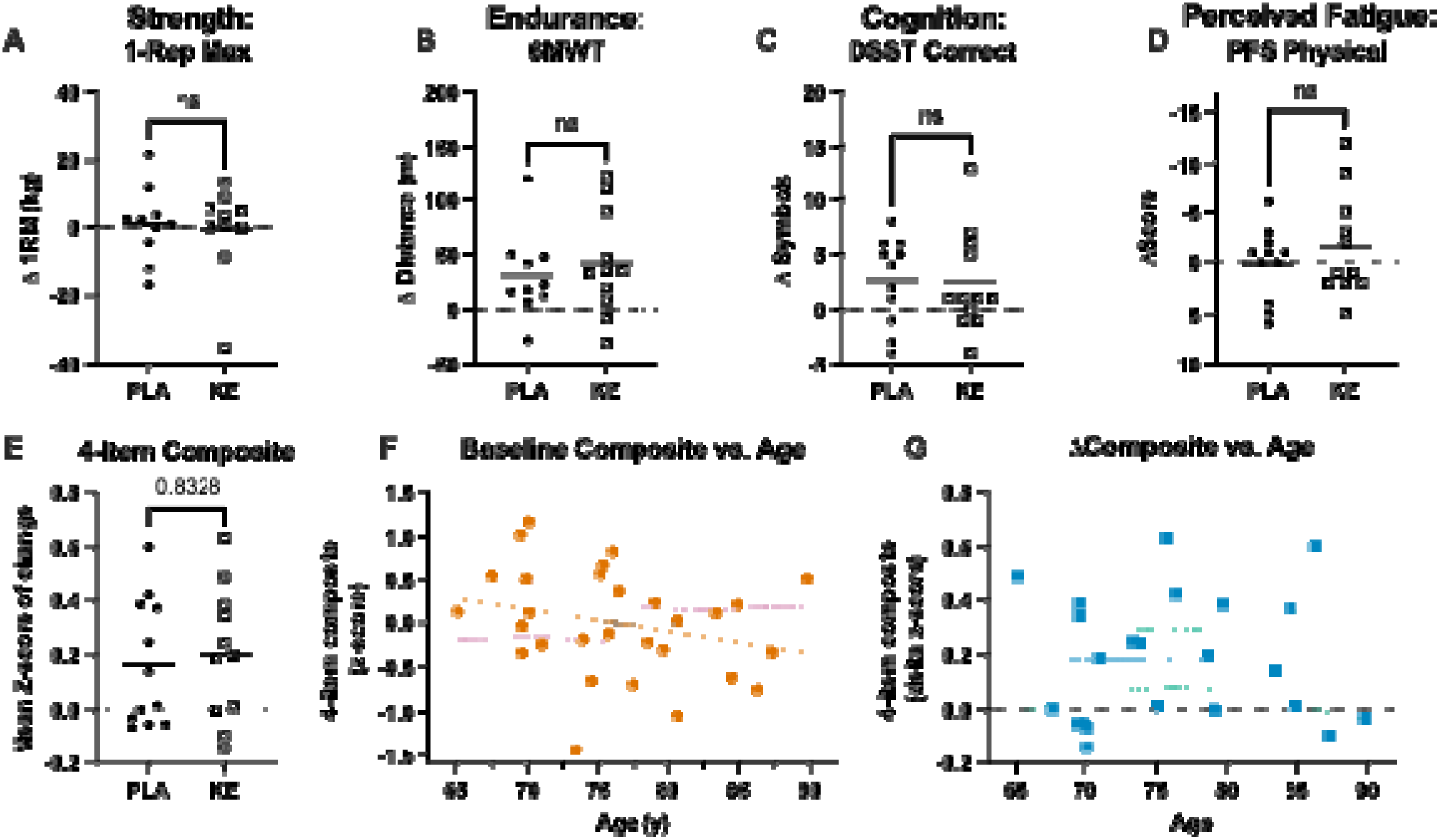
Change in four functional outcomes from pre- to post- 12 weeks of daily consumption of ketone ester or placebo in n = 23 healthy older adult. **A:** 1 repmax leg press, **B:** six minute walk test, **C:** digit symbol substitution task, **D** Pittsburgh Fatigability Scale. Change in the composite score resulting from the combination of these four outcomes (**E**), and characteristics of the composite score (**F,G**). **Abreviations**: KE, ketone ester; PLA, placebo; 6MWT, six minute walk test; DSST, digit symbol substitution task, PFS, Pittsburgh Fatigability Scale. Y-axes are oriented so higher is better in all panels.

### Physical function

There were no signigicant differences between intervention groups in the longitudinal change in the remaining physical function outcomes (**Figure 3 A-C**): grip strength (KE = -1 (4.8) kg; PLA = 0.6 (6.0) kg, p = 0.728), Short Physical Performance Battery (KE = 0.0 (0.4); PLA = 0.4 (0.5), p = 0.197) or leg press reps to fatigue at 70% of maximal weight (KE = -0.7 (7.3); PLA = -1.1 (5.2), p =0.982).

**Figure 3:**
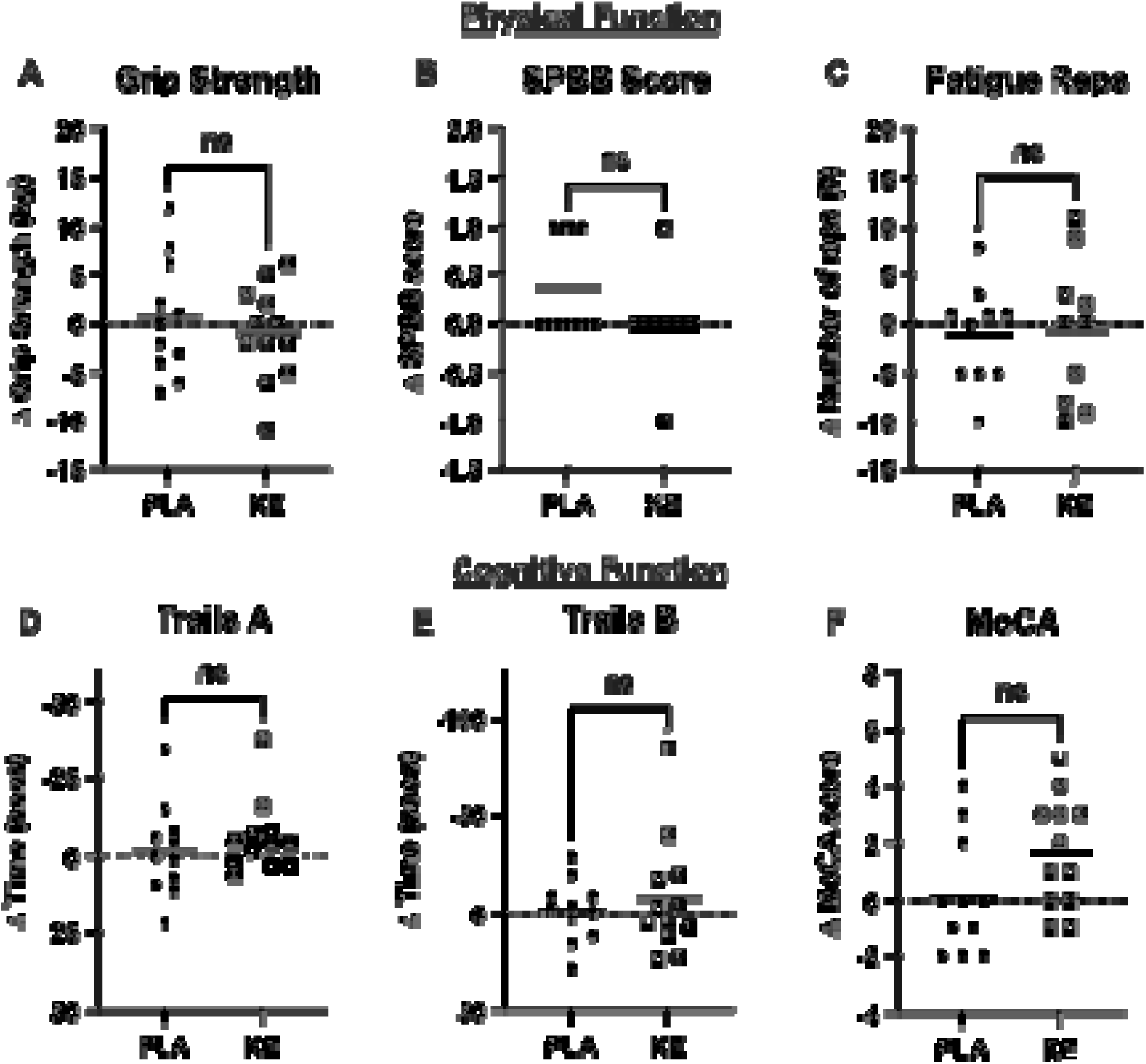
Changes in physical and cognitive function outcomes from pre- to post- 12 weeks of daily consumption of ketone ester or placebo in n = 23 healthy older adults. **A:** grip strength, **B:** short physical performance battery, **C:** leg press reps to fatigue at 70% maximal weight, **D:** Trails A, **E:** Trails B, **F:** Montreal Cognitive Assessment. **Abbreviations**: KE, ketone ester; PLA, placebo; MoCA, Montreal Cognitive Assessment; SPBB, Short Physical Performance Battery. Y-axes are oriented so higher is better in all panels.

### Cognitive function

There were no significant differences between intervention groups in the longitudinal change in the remaining cognitive function outcomes (**Figure 3D-E**): Trails A (KE = -5.8 (11.8) s; PLA = -1.6 (12.1) s, p =0.422), Trails B (KE = -7.8 (30.5) s; PLA = -1.4 (16.3) s, p > 0.999) or Montreal Cognitive Assessment (KE = 1.7 (2.0); PLA = 0.1 (2.1), p = 0.068).

### Quality of life

There were no differences between study groups in the longitudinal change in any of the subdomains or global summary scores (where appropriate) of the Profile of Mood States, SF-36, Geriatric Depression Scale, Sexual Quality of Life questionnaire, or the Pittsburgh Fatigability Scale (**Table 1**). Sleep, assessed by the Pittsburgh Sleep Quality Index (PSQI) demonstrated a worsening in subjective sleep efficiency in the KE group (KE = 0.8 (1.6); PLA = -0.5 (1.1), p = 0.058) and a significant worsening of the global PSQI index in the KE group (KE = 1.8 (2.6); PLA = -0.3 (2.3), p = 0.047) (**Table 1**).

**Table 1:**
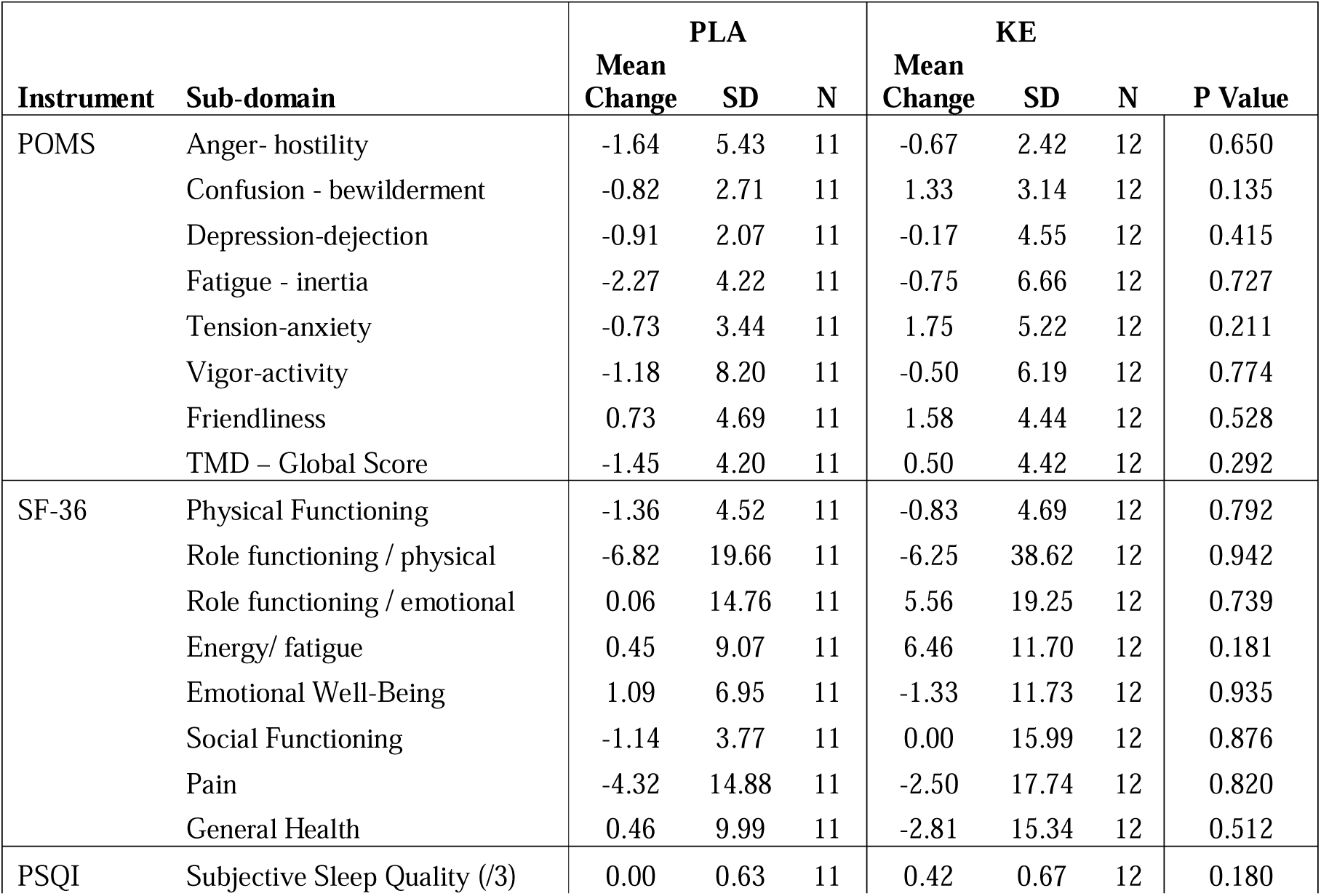
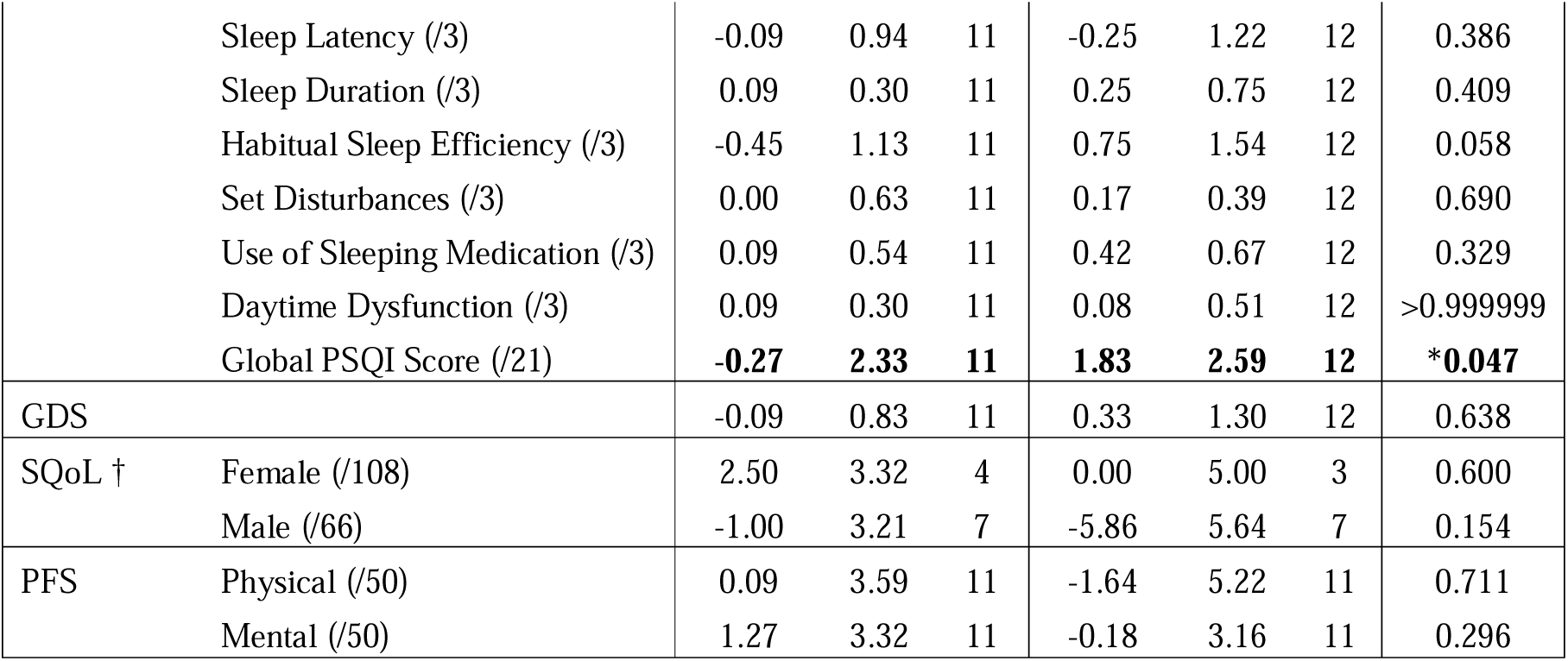
Summary of the pre- post- intervention change in quality of life outcomes in. **Abbreviations:** GDS, Geriatric Depression Scale; PFS, Pittsburgh Fatigability Questionnaire;POMS, Profile of Mood States; PSQI, Pittsburgh Sleep Quality Index; SD, Standard Deviation; SF-36, Short Form 36; SQoL, Sexual Quality of Life. † = some subjects declined to complete the SQoL, * = p < 0.05.

| Instrument | Sub-domain | PLA |  |  | KE |  |  | P Value |
| --- | --- | --- | --- | --- | --- | --- | --- | --- |
|  |  | Mean Change | SD | N | Mean Change | SD | N |  |
| POMS | Anger- hostility | -1.64 | 5.43 | 11 | -0.67 | 2.42 | 12 | 0.650 |
|  | Confusion - bewilderment | -0.82 | 2.71 | 11 | 1.33 | 3.14 | 12 | 0.135 |
|  | Depression-dejection | -0.91 | 2.07 | 11 | -0.17 | 4.55 | 12 | 0.415 |
|  | Fatigue - inertia | -2.27 | 4.22 | 11 | -0.75 | 6.66 | 12 | 0.727 |
|  | Tension-anxiety | -0.73 | 3.44 | 11 | 1.75 | 5.22 | 12 | 0.211 |
|  | Vigor-activity | -1.18 | 8.20 | 11 | -0.50 | 6.19 | 12 | 0.774 |
|  | Friendliness | 0.73 | 4.69 | 11 | 1.58 | 4.44 | 12 | 0.528 |
|  | TMD – Global Score | -1.45 | 4.20 | 11 | 0.50 | 4.42 | 12 | 0.292 |
| SF-36 | Physical Functioning | -1.36 | 4.52 | 11 | -0.83 | 4.69 | 12 | 0.792 |
|  | Role functioning / physical | -6.82 | 19.66 | 11 | -6.25 | 38.62 | 12 | 0.942 |
|  | Role functioning / emotional | 0.06 | 14.76 | 11 | 5.56 | 19.25 | 12 | 0.739 |
|  | Energy/ fatigue | 0.45 | 9.07 | 11 | 6.46 | 11.70 | 12 | 0.181 |
|  | Emotional Well-Being | 1.09 | 6.95 | 11 | -1.33 | 11.73 | 12 | 0.935 |
|  | Social Functioning | -1.14 | 3.77 | 11 | 0.00 | 15.99 | 12 | 0.876 |
|  | Pain | -4.32 | 14.88 | 11 | -2.50 | 17.74 | 12 | 0.820 |
|  | General Health | 0.46 | 9.99 | 11 | -2.81 | 15.34 | 12 | 0.512 |
| PSQI | Subjective Sleep Quality (/3) | 0.00 | 0.63 | 11 | 0.42 | 0.67 | 12 | 0.180 |
|  | Sleep Latency (/3) | -0.09 | 0.94 | 11 | -0.25 | 1.22 | 12 | 0.386 |
|  | Sleep Duration (/3) | 0.09 | 0.30 | 11 | 0.25 | 0.75 | 12 | 0.409 |
|  | Habitual Sleep Efficiency (/3) | -0.45 | 1.13 | 11 | 0.75 | 1.54 | 12 | 0.058 |
|  | Set Disturbances (/3) | 0.00 | 0.63 | 11 | 0.17 | 0.39 | 12 | 0.690 |
|  | Use of Sleeping Medication (/3) | 0.09 | 0.54 | 11 | 0.42 | 0.67 | 12 | 0.329 |
|  | Daytime Dysfunction (/3) | 0.09 | 0.30 | 11 | 0.08 | 0.51 | 12 | >0.999999 |
|  | Global PSQI Score (/21) | **-0.27** | **2.33** | **11** | **1.83** | **2.59** | **12** | **\*0.047** |
| GDS |  | -0.09 | 0.83 | 11 | 0.33 | 1.30 | 12 | 0.638 |
| SQoL † | Female (/108) | 2.50 | 3.32 | 4 | 0.00 | 5.00 | 3 | 0.600 |
|  | Male (/66) | -1.00 | 3.21 | 7 | -5.86 | 5.64 | 7 | 0.154 |
| PFS | Physical (/50) | 0.09 | 3.59 | 11 | -1.64 | 5.22 | 11 | 0.711 |
|  | Mental (/50) | 1.27 | 3.32 | 11 | -0.18 | 3.16 | 11 | 0.296 |

### Health and Fitness Tracker

We observed a qualitative difference in sleeping heart rate (SHR, the mean heart rate during one sleep period; see **Methods**) trends between the KE and PLA groups, with the KE arm showing a sustained elevation in SHR (**Figure 4A**). In contrast to the PSQI data indicating lower subjective sleep quality with the KE, we did not see any differences in between groups trend in sleep minutes (**Figure 4B**) or efficiency (**Supplemental Figures**). There were no differences in steps (**Figure 4C**) or total active minutes (**Figure 4D, Supplemental Figures**) between groups. For all measures, the small effect size of the cohort and the high inter-individual variability prohibited any meaningful quantification of the difference between the two arms.

**Figure 4:**
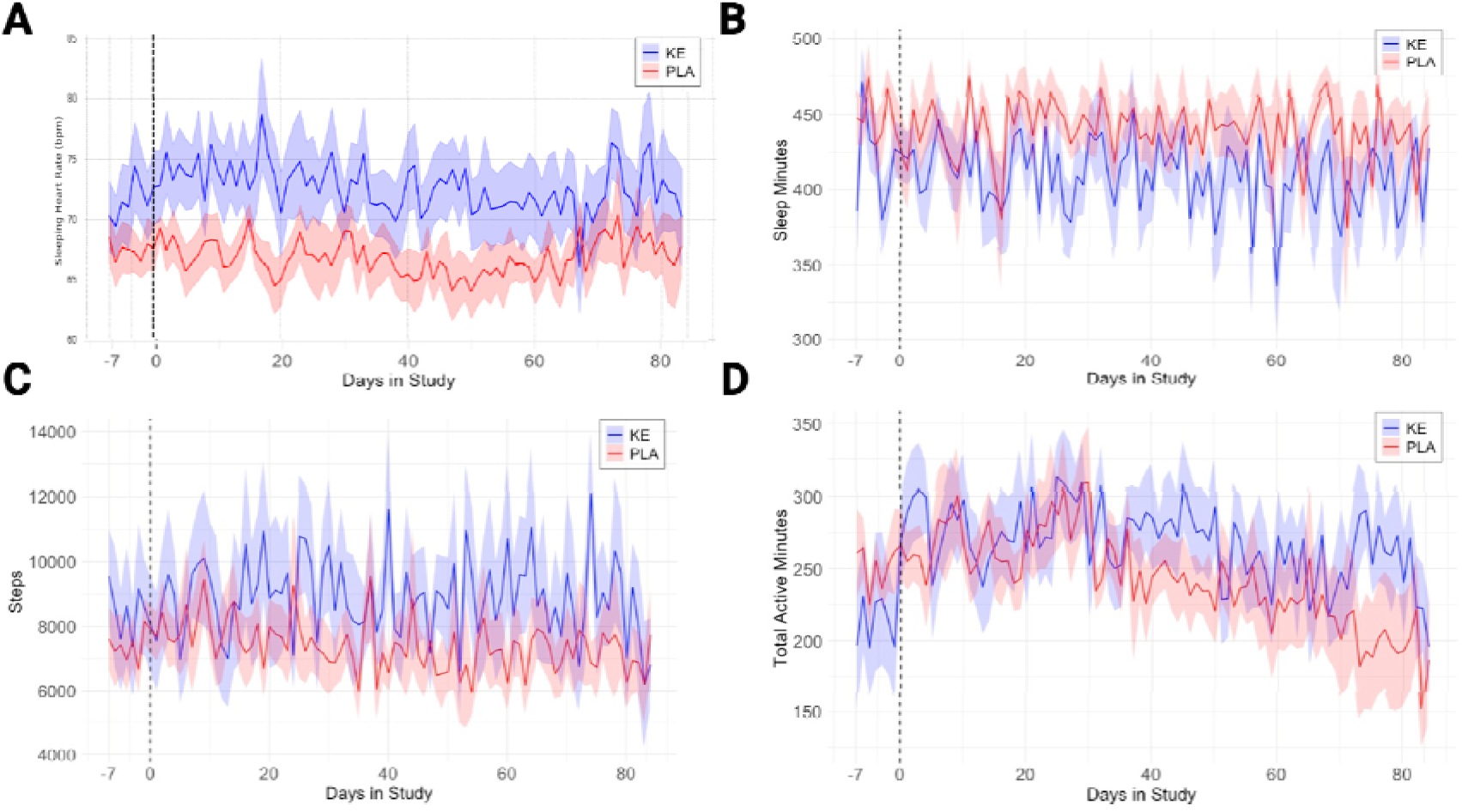
Data from wearable health and fitness trackers worn by study participants who completed the 12 week protocol, showing mean and standard error for A) sleeping heart rate, B) sleep minutes, C) daily steps, D) total active minutes for 7 days before baseline visit, and for the remaining 12 weeks of the study. Abbreviations: KE, ketone ester; PLA, placebo.

## Discussion

The main finding of this exploratory analysis of the effects of 12 weeks of KE consumption on functional outcomes in a pilot study of healthy older adults was that, in contrast to our hypothesis, there were no statistically significant effects of the KE intervention on a composite score designed to capture the vigor-frailty continuum, or on individual items scoring physical or cognitive function, activity, resting heart rate or quality of life. However, this tolerability-focused pilot randomized controlled trial was not powered for functional outcomes, and enrolled non-frail, healthy older adults. As an example of an early-stage geroscience clinical trial, we discuss the rationale for intervention-specific outcome measures as well as the selection of common measures that broadly capture function in aging and represent a common toolbox for geroscience clinical trials (39, 40).

Ketone bodies are expected to benefit physical function via multiple distinct mechanisms. Firstly, ketones might act as an alternative energy substrate that directly improved working muscle efficiency (41). Secondly, there is a known acute effect of exogenous ketones on cardiac output and myocardial blood flow (42), which may indirectly facilitate physical function. Thirdly, ketones are known to be anticatabolic and may increase muscle protein synthesis and decrease muscle protein breakdown in the context of inflammatory stress (43, 44), feasibly such as that seen during age-related or frailty-related inflammation, which could preserve or increase muscle mass and function with longer term use. Some, but not all, studies that administered exogenous ketones to young athletes immediately prior to endurance exercise found functional improvements (24, 25). The only study of physical function with longer term exogenous ketone consumption found that KE could mitigate the performance decline and hormonal shifts triggered by ‘over-reaching’ endurance training in young adults (45). Clinical evidence for a muscle sparing function of exogenous ketones is limited. Two studies used ketone infusions in healthy young men and found attenuated leucine oxidation and increased muscle protein synthesis (43), and a decreased muscle protein breakdown in the context of an inflammatory stressor (44). A further study gave a ketone ester drink and found lowered post exercise AMPK phosphorylation and higher mTORC1 activation, suggesting greater protein synthetic potential (46). None of these studies investigated functional changes in muscle strength.

While this study is part of a program to test the long-term effects of chronic daily use of KE in frailty, these data from athletes suggest that other approaches might be fruitful to pursue in parallel. For example, the greatest performance gains might be acutely during ketosis, perhaps relevant to scheduling the use of ketones as rehabilitation or exercise adjuvants. A concurrent stressor might increase the effects from ketones. Heart failure or other medical problems associated with energetic stress might be seen as examples of such stressors, but a more benign stressor might be resistance training. A parallel can be drawn with dietary protein supplementation, which alone does not consistently improve muscle mass and function in older adults (47), but increases the efficacy of resistance training compared to a placebo (48, 49). Future work could address combinations of ketones, protein and resistance exercise to determine if there any synergistic effects of these strategies acutely and long-term.

The rationale for an effect of exogenous ketones on cognitive function is also multifactorial. Firstly, ketones act as an energy source in the brain and can mitigate the deficit in brain energetic needs that arises during age-related declines in glucose metabolism (50). Secondly, ketones increase brain blood flow which would improve delivery of substrates and oxygen (27, 51). Thirdly, ketones can trigger the release of neurotrophins, particularly brain derived neurotrophic factor (BDNF) (52). Key clinical examples include a 12-month study of two 15g daily servings of a ketogenic medium chain triglyceride, which found improved brain energy metabolism and cognition in adults with mild cognitive impairment (53), and a 14-day study of three 12 g daily servings of a ketone ester which found improved cerebral blood flow and elements of cognition in obese adults (27). As there is a strong biological mechanism and both preclinical and clinical support for a neurocognitive effect of exogenous ketones, outcome measures relating to brain physiology and function will remain of keen interest in future work.

Whilst they are many steps removed from the underlying biological mechanisms and likely are influenced by multiple mechanisms, subjective self-assessment of quality of life is increasingly recognized as an important outcome in interventional geroscience trials that can distinguish functionally healthy and unhealthy aging (54). To this end, we included a range of validated quality of life questionnaires, notably the SF-36, which has been proposed as a leading quality of life assessment tool (54).

We did observe a decline in subjective sleep efficiency and overall sleep score in the KE population during the study, although there were no matching trends in sleep time or efficiency measured by our health and fitness tracker data. It is unclear if the trend for increased SHR in the KE group could have contributed to lower sleep quality; acutely increased heart rate following administration of exogenous ketones has been reported in some studies (29, 51, 55), but heart rate returns to basal values when plasma BHB falls (27, 56), although no data from a continuous wearble exists to our knowlege. Only one published study has investigated the interaction between exogenous ketones and sleep, finding that consumption of a ketone ester before bed, after high intensity exercise, improved sleep quality in young athletes (57). Our results indicated the opposite effect, although many contextual elements are different between studies, including KE timing, study population, and the addition of exercise as a stressor. Sleep is a critical component of health (58), and sleep quality generally declines with age (59), therefore future studies using KE in older adults should continue to monitor sleep quality, along with other quality of life domains to definitively elucidate any effects of KE.

We chose to explore the sensitivity of a composite score for vigor-frailty as it is increasingly appreciated that many gerotherapeutic strategies, such as BO-BD, could have pleiotropic and potentially modest effects across a variety of individual organ systems (e.g., muscle strength, endurance, cognitive function) but have a clinically important overall effect on the whole person due to these integrated, potentially synergistic, multi-system effects (60). It should be noted that the use of a composite is not without downsides, as a study using a composite score might fail to capture substantial changes within just one domain if not statistically powered for that endpoint alone. Composite outcomes are increasingly used in geroscience focused studies and range from composites of death or disease onset (e.g., Targeting Aging with Metformin - TAME), of blood and clinical biomarkers (e.g., in SGLT-2 trials) or of functional endpoints (e.g., Intrinsic Capacity). Composite cardiovascular outcomes (e.g. Major Adverse Cardiovascular Events, MACE) are widely used in clinical trials (61). Given the frailty expertise of the SOMMA investigators, and the size and richness of the SOMMA dataset, we chose to adapt the SOMMA-derived composite vigor-to-frailty outcome using leg press weight rather than leg power, and 6-minute walk test distance instead of a direct VO_2_ max measurement as a measure of endurance, as this 6-minute walk is commonly used to estimate VO_2_ max (62). As we expected, our baseline composite score showed a trend of decreasing with age even in our small and functionally independent population. We did not see any difference in composite score between intervention groups, and importantly we did not see an age-effect on change in composite score. Overall, the data from this cohort provided an interesting opportunity to explore the implementation of a frailty focused composite outcome, which might have an increased likelihood of detecting any KE driven effects in future studies.

The strengths of this study include the free-living, pragmatic-inspired design (e.g. participants were instructed to maintain their usual diet and exercise habits) being highly relevant for future uses of KE as a geroscience intervention, the high adherence observed, the equal enrollment of men and women, the average age (76 y) being well over the lower limit (65 y), and the close matching achieved between the KE and PLA beverage, although the findings should be interpreted with the context that the lipid-based PLA was not truly ‘inert’ and thus does not offer a comparison to ‘no intervention’.

There were several limitations of this study, many of which were inherent to its design as a “first in older adults” early-stage geroscience clinical trial (60), and that we plan to address in our follow-up study. The primary goals of this pilot study were to fill key gaps in kinetics, tolerability, and safety in older adults that would unlock the ability to design function-focused clinical trials with KE in older adults. The 12-week study duration was selected conservatively due to the longest prior study of any KE in any population being only 4 weeks, and the unknown feasibility and safety of the intervention in older adults. However, 12 weeks is relatively short to expect detectable changes in functional outcomes which often take months or years to manifest. Furthermore, whilst the 25 g once daily serving size was selected based on the previous studies of BO-BD in young adults, and the absence of any data on any dose of KE in older adults, studies of other KE in young adults have used servings of up to 75 g daily split across three doses. Further increasing blood ketone concentrations or overall exposure time with larger or more frequent dosing may increase response. It is also important to note that several types of exogenous ketones and KEs exist, with known differences in physical characteristics and possibly also functional effects, therefore our results using BO-BD may not apply to all types of exogenous ketones. In addition, the optimal outcome measures for early-stage geroscience clinical trials have not been defined. There is clear advantage to using measures that are well validated and commonly used in large longitudinal studies, though some of these may be best suited to much longer multi-year studies. Many of these outcomes also have considerable inter-individual variability across testing occasions, as well as training effects that limit repetition. Novel functional biomarkers may help bridge the gap between target engagement or kinetic outcomes (e.g. blood BHB levels) and clinically relevant, patient-centered, long-term functional outcomes. Notably, we chose to conduct all functional testing on days without KE consumption to focus on stable, sustained effects of KE rather than transient performance associated with acute ketosis (and to avoid confounding from the sequencing of study activities with respect to the peak of post-ingestion blood BHB concentrations). However, it is possible that ongoing ketosis is required for performance gains. Specific investigation will be required to answer this important question. Finally, the small sample size and relatively healthy and fit population, both determined by the primary study goals, were major limitations in our ability to detect a difference in these exploratory functional outcomes. We expect that a larger sample size, and a study population selected for increased vulnerability or with existing mild functional limitations would increase the likelihood of a detectable positive effect.

In conclusion, consuming the KE, BO-BD, daily for 12 weeks did not impact exploratory quality of life, physical or cognitive functional outcomes in this pilot cohort of healthy older adults. Future work using a larger cohort of pre-frail and early frail older adults will seek to definitively test if the hypothesized benefits of exogenous ketones will be detectable against a functionally limited baseline.

## Abbreviations

1-RM: 1 Repetition Maximum
ADL: Activities of Daily Living
AEs: Adverse Events
BDO: (R)-1,3-butanediol
BH-BD: Bis-hexanoyl (R)-1,3-butanediol
ΒHB: Beta-hydroxybutyrate
BMI: Body Mass Index
BO-BD: Bis-octanoyl (R)-1,3-butanediol
BTQ: Beverage Tolerability Questionnaire
CSHA: Canadian Study of Health and Aging
CRF: Case Report Form
DSST: Digit Symbol Substitution Test
IADL: Instrumental Activities of Daily Living
ITT: Intention to treat
KEs: Ketone Esters
MoCA: Montreal Cognitive Assessment
PLA: Placebo
PP: Per protocol
QoL: Quality of Life
SASP: Senescence-associated Secretory Phenotype
SPPB: Short Physical Performance Battery

## Sources of Funding, Author Declarations and Conflict Management

Funding for the study was provided by philanthropic donations from Dr. James B. Johnson and from members of the Buck Institute Impact Circle. Dr. Johnson assisted with conceptualization of the study and reviewed this manuscript but has no further role in study design, management, data collection, analysis, interpretation of data, decision to submit publications, or writing of publications. The Buck Institute Impact Circle had no role in conceptualization, study design, management, data collection, analysis, interpretation of data, decision to submit publications, review, or writing of publications.

Dr. Newman’s participation in the study was supported by Buck Institute institutional funds. Dr. Brianna Stubbs’ participation in the study was supported by the NIH (NIA) under award number K01AG078125.

The KE intervention was provided gratis by BHB Therapeutics Ltd (Ireland). BHB Therapeutics also arranged for manufacture of the matched placebo, paid for by study funds. BHB Therapeutics (Ireland) markets formulated KE beverages to consumers. BHB Therapeutics (Ireland) provided no funding for the study, and had no role in the design, management, data collection, analysis, interpretation of data, decision to submit publications, or writing of publications.

## Author Declarations and Conflict Management

The Buck Institute holds shares in BHB Therapeutics (Ireland) and Selah Therapeutics. B.J.S. has stock in H.V.M.N Inc, and stock options in Selah Therapeutics Ltd, BHB Therapeutics (Ireland) Ltd., and Juvenescence Ltd. J.C.N. has stock options in Selah Therapeutics Ltd and BHB Therapeutics (Ireland) Ltd. J.C.N and B.J.S. are inventors on patents related to the use of ketone bodies that are assigned to The Buck Institute. Individual and institutional conflict management plans were developed and approved by the Buck Institute and submitted to the reviewing IRB. Actions and decisions important to participant safety and study integrity were carried out by ‘honest brokers’ with no potential financial conflict. Participant consent was obtained by licensed registered nurses (L.A and W.S.M) who have no financial conflict. Decisions on participant enrollment, continuation, and were made by independent medical officers (J.M and M.Y) unaffiliated with Buck Institute and with no financial conflict. All other authors have no conflicts to report.

## Author Contributions

Conceptualization, J.C.N, B.J.S, J.B.J; methodology, J.C.N, B.J.S, J.B.J; investigation, B.J.S, E.B.S, C.S, S.R.D, S.P, W.S-M, L.A, M.Y, J.M; data curation, B.J.S, E.B.S, C.S, S.R.D, S.P, J.K; formal analysis, B.J.S, E.B.S, J.K, J.T.Y, A.F; writing—original draft preparation, B.J.S, E.B.S; writing—review and editing, all; visualization, B.J.S, E.B.S; project administration, B.J.S, J.C.N, T.G; supervision: B.J.S, T.G, J.C.N; funding acquisition, B.J.S, J.C.N, J.B.J; All authors have read and agreed to the published version of the manuscript.

## Data Availability Statement

The data presented here may be available upon reasonable request from the corresponding author and in accordance with intellectual property considerations.

## Supplementary Information – Full Inclusion and Exclusion Criteria

### Inclusion Criteria

1. Subject is greater than or equal to 65 years of age, inclusive at Visit 1.
2. Subject has a BMI 18.5-34.9 kg/m^2^ (inclusive) at Visit 1.
3. Subject is willing and able to comply with all study procedures including randomization into any of the experimental groups, maintenance of habitual dietary intake, exercise and medication and supplement use, blood draws and the following prior to test visits: fasting (≥10 h; water only), no alcohol (≥ 10 h), no cannabis products (≥10 h) and no exercise (≥ 10 h).
4. Subject has no health conditions that would prevent them from fulfilling the study requirements as judged by the Clinical Investigator on the basis of medical history and routine laboratory test results.
5. Subject understands the study procedures and signs forms providing informed consent to participate in the study.

### Exclusion Criteria

2. Subject is non ambulatory
3. Subject has a CSHA clinical frailty score > 5
4. Subject requires assistance with any activity of daily living, excluding continence
5. Subject lives in an institutional setting (skilled nursing facility or residential care facility for the elderly).
6. Subject is a female who has not passed menopause.
7. Subject is unable to converse in English
8. Subject is unable to provide informed consent due to cognitive impairment or insufficient English language comprehension
9. Subject has been hospitalized within 30 days of Visit 1, 2 or 3.
10. Subject has an abnormal laboratory test result(s) of clinical importance, indicating unstable chronic disease of major organ dysfunction, at Visit 1, at the discretion of the Medical Officer. One re-test will be allowed on a separate day prior to Visit 2, for subjects with abnormal laboratory test results.
11. Subject has a history or presence of uncontrolled and/or clinically active pulmonary, cardiac (e.g. >= New York Heart Association class III), hepatic, renal, endocrine (including type 1 diabetes), hematologic, immunologic, neurologic (e.g., Alzheimer’s or Parkinson’s diseases), psychiatric (including unstable depression and/or anxiety disorders) or biliary disorders. Stable chronic disease is not an exclusion criterion unless specified.
12. Subject has a clinically important gastrointestinal condition that would potentially interfere with the evaluation of the study beverage [e.g., inflammatory bowel disease, irritable bowel syndrome, chronic constipation, severe constipation (in the opinion of the Clinical Investigator), history of frequent diarrhea, history of surgery for weight loss, gastroparesis, systemic disease that might affect gut motility according to the Investigator, reflux requiring daily medication, history of gastrointestinal ulcers or bleeding, and/or clinically important lactose intolerance].
13. Subject has a history of alcohol or substance abuse.
14. Subject is consistently using prescriptive or over-the counter medications where alcohol is a contraindication at the discretion of the Investigator.
15. Subject has a known allergy, intolerance, or sensitivity to any of the ingredients in the study beverages, including soy and milk protein.
16. Subject has uncontrolled hypertension (systolic blood pressure ≥140 mm Hg or diastolic blood pressure ≥90 mm Hg) as defined by the blood pressure measured at Visit 1. One re-test will be allowed on a separate day before Visit 2, for subjects with abnormal blood pressure.
17. Subject is undergoing treatment or active surveillance for cancer, or has been diagnosed with cancer in the prior two years, except for non-melanoma skin cancer.
18. Subject has recently used antibiotics within 30 days of Visit 1, 2 or 3.
19. Subject has extreme dietary habits (e.g., intermittent fasting or time restricted eating, Atkins diet, vegan, very high protein/low carbohydrate or has used weight-loss medications (including over-the-counter medications and/or supplements) or programs within 30 days of Visit 1, 2 or 3.
20. Subject has used medications (over-the-counter or prescription) known to influence gastrointestinal function including, but not limited to, opioids, weight loss medications, antidiarrheals, and antispasmodics) within 30 days of Visit 1, 2 or 3.
21. Subject has used ketone supplements (ketone salts or esters, and medium chain triglycerides [MCT]) within 30 days of Visit 1, 2 or 3.
22. Subject has unstable use of thyroid, antihypertensive, antidepressant, or statin medications within 30 days of Visit 1, 2 or 3.
23. Subject has a condition the Clinical Investigator believes would interfere with their ability to provide informed consent, comply with the study protocol, which might confound the interpretation of the study results, or put the subject at undue risk.
24. Subject works nights or shifts that means it is not possible to maintain a consistent meal schedule during the study.
25. Subject is not permitted to visit the Buck Institute campus, for example due to inability to confirm COVID-19 vaccination status.
26. Subject does not have a Bluetooth enabled smartphone.
27. Subject does not have access to the internet

### Excluded Medications/Supplements/Products

Use of thyroid hormone therapy, statins, antihypertensives, antidepressants, constipation medications should be **<u>stable</u>** for the 30 days prior to Visit 1, 2 or 3. Additionally, use of any antibiotic therapy is not permitted within 30 days of Visit 1, 2 or 3 and throughout the study period. Subjects should not use opioids, weight loss medications, antidiarrheals, antispasmodics, ketone supplements (including MCT oil), or other medications (over-the-counter or prescription) or dietary supplements known to alter gastrointestinal function within 30 days of Visit 1, 2 or 3 and throughout the study period, with the exception of stable use of constipation medications and supplements.

Should a subject require any of these medications or supplements, the study staff should consult with the Investigator to discuss the subject’s continued participation in the trial. At the discretion of the Medical Officer in consultation with the Investigator, subjects may suspend consumption of the Study Product and completion of the Study Log for up to 10 days and re-start the protocol at the point of suspension.

---

**Supplementary Figure 1.**
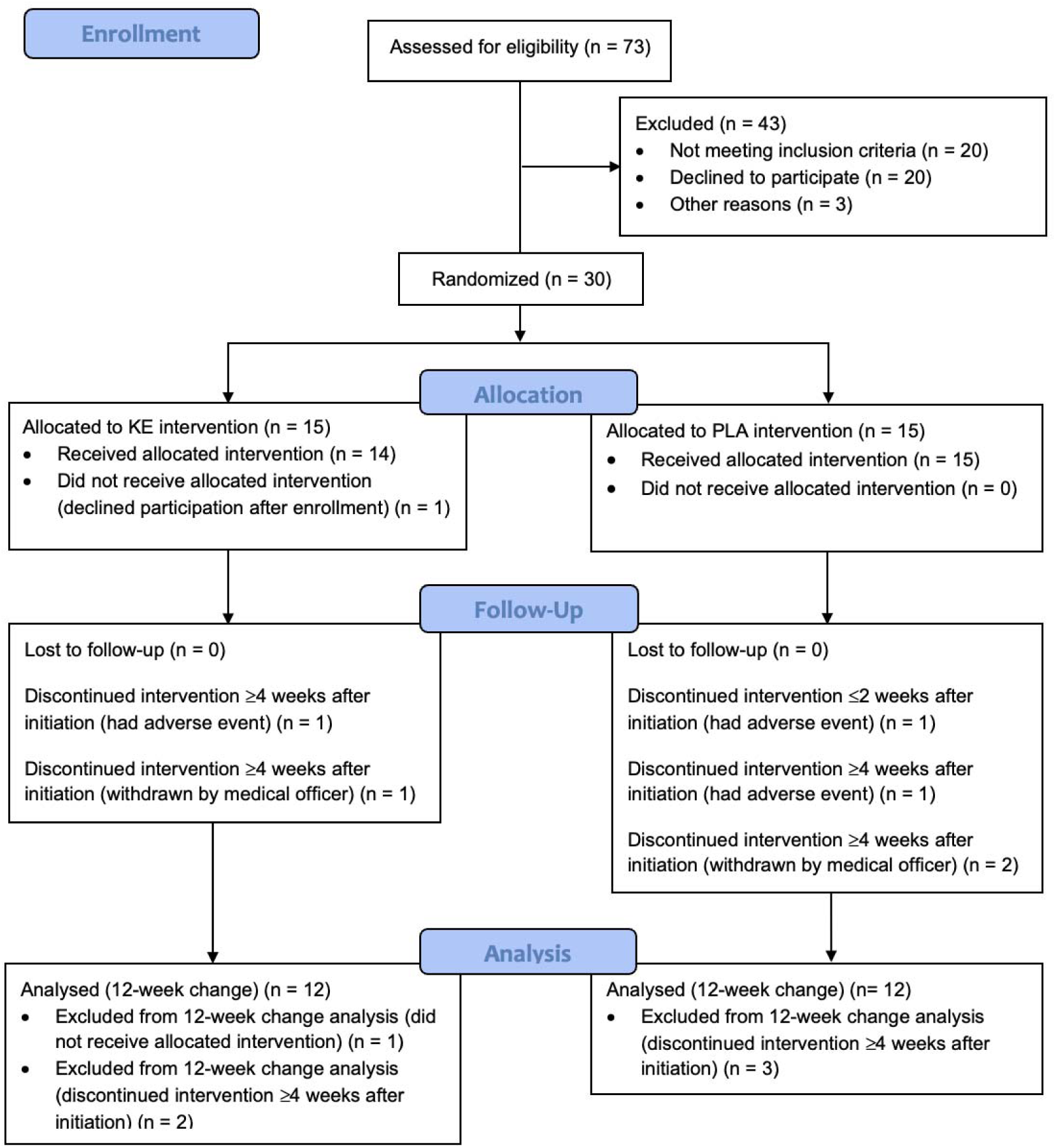

**Supplementary Table 1:**
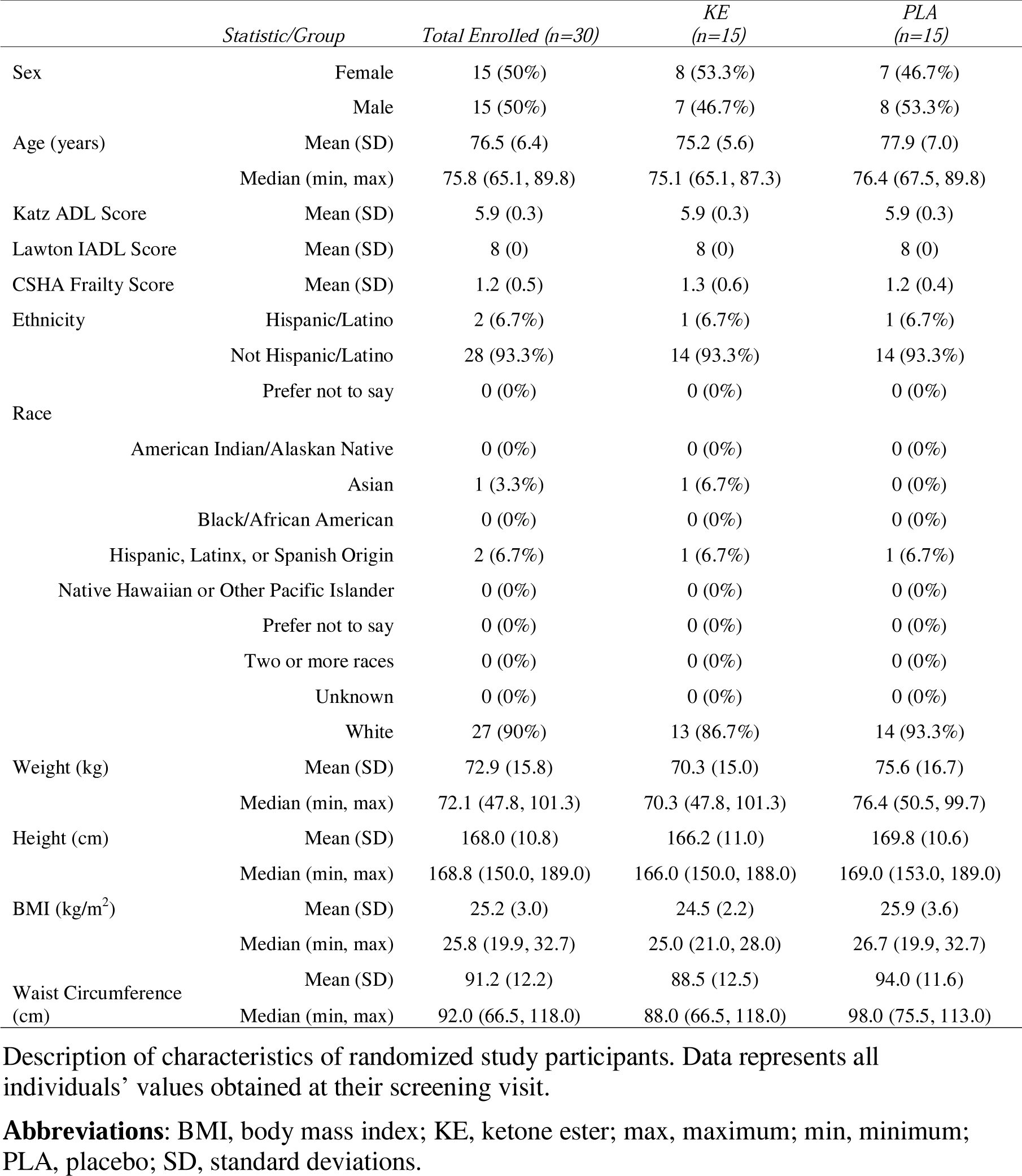
Study participant characteristics.

**Supplementary Table 2.** Nutritional Information. Nutritional information for the KE and PLA beverages. **Abbreviations:** BO-BD, bis-octanoyl (R)-1,3 butanediol; KE, ketone ester; PLA, placebo

|  | KE | PLA |
| --- | --- | --- |
| BO-BD (g) | 25 | 0 |
| Energy (kcal) | 240 | 246 |
| Carbohydrate (g) | 2 | 2 |
| Fat (g) | 0.5 | 25 |
| Protein (g) | 2 | 2 |

